# Incorporation of Visit-to-Visit Blood Pressure Variability into Cardiovascular Disease Risk Prediction

**DOI:** 10.64898/2026.03.03.26347482

**Authors:** Mifetika Lukitasari, Nickson Ning, Siaw-Teng Liaw, Bin Jalaludin, Joel Rhee, Jitendra Jonnagaddala

## Abstract

**BACKGROUND:** Visit-to-visit blood pressure variability (VVV BPV) is an important yet underutilised risk factor for cardiovascular disease (CVD) risk prediction. Incorporating VVV BPV in the model predicting CVD could improve its performance. This study aims to incorporate VVV BPV into a CVD risk prediction model and to evaluate its performance by comparing the discrimination and calibration of models using a single BP measurement versus those incorporating VVV BPV

**METHODS:** This prospective cohort study included data from the electronic practice-based research network (ePBRN) in Southwestern Sydney, focusing on patients aged 18-55 years with at least five BP readings, excluding those with incomplete data or no follow-up after 55. VVV BPV measured by standard deviation (SD) and coefficient of variation (CV). The main outcome is the first occurrence of CVD. We developed the models using Cox proportional hazards regression with 10-fold cross-validation on all imputed datasets. Model performance was evaluated for discrimination and calibration. Discrimination was assessed using Harrell’s C-index and time-varying AUC for five-year CVD prediction. Calibration was assessed using calibration slopes and Brier scores, which were also evaluated annually.

**RESULTS:** The study involved 3,065 patients, with 45.41% women. Incorporating VVV BPV improved the prediction of CVD risk in people aged 55 years. The model with a single systolic blood pressure (SBP) measurement had a Harrel C-Index of 0.716 (95% CI: 0.658 - 0.775), while those using SD and CV scored higher at 0.833 (95% CI: 0.804 - 0.862) and 0.837 (95% CI: 0.810 - 0.864), respectively. Five years’ AUC for SBP was 0.852 (95% CI: 0.820 – 0.885) for SD and 0.856 (95% CI: 0.824 – 0.888) for CV. In contrast, the single SBP model had a lower AUC of 0.757 (95% CI: 0.700 – 0.815). No significant difference was observed in calibration slopes and Brier scores between the model using single BP and VVV BPV.

**CONCLUSIONS:** This study developed a model for CVD risk estimation using VVV BPV instead of a single blood pressure measurement. Replacing a single BP measure with VVV BPV significantly enhanced the model’s predictive accuracy.

## Introduction

Cardiovascular disease (CVD) is a leading cause of morbidity and mortality among older adults. Current prevention guidelines for people without pre-existing CVD recommend stratifying patients’ 5- or 10-year CVD risk using established prognostic models developed from a specific population^1–5^ The Framingham Risk Score (FRS) from the USA is one of the earliest and most widely recognised tools for estimating CVD risk and has been adapted for use in various populations across different countries. The 2003 Australian CVD risk assessment model, derived from the FRS, has shown limitations, tending to overestimate the risk in the general Australian population^6^ while underestimating the risk among Aboriginal and Torres Strait Islander people.^7^

High blood pressure (BP) is a well-known risk factor for CVD. However, recent attention has shifted from relying on a single BP measurement to considering repeated BP measurements over time.^8–11^ Most existing CVD risk prediction models incorporate only a single BP measurement, despite evidence that BP fluctuates substantially between clinical visits. A previous study suggested that Visit-to-visit blood pressure variability (VVV BPV) is common among primary care patients.^12^ Our previous work identified cut-off values for the standard deviation (SD) (one of the most popular BPV metrics) for predicting CVD within five years. However, no prior study has incorporated BPV using empirically derived cut-off values into CVD risk prediction models.

Clinical data contained in electronic health records (EHRs) hold substantial potential for disease risk prediction and personalised treatment; however, current CVD risk prediction models do not fully leverage longitudinal data, such as repeated BP measurements collected during routine clinic visits. Several studies have recommended the use of longitudinal data to enhance the discrimination of CVD risk across different populations.^13–17^ A prior study developing and validating the QRISK3 model attempted to incorporate VVV systolic BPV by adding the SD of systolic blood pressure (SBP) to the most recent SBP value. However, this modification did not improve the model’s discrimination or calibration.^18^ In contrast, another study involving patients with diabetes mellitus revealed that incorporating variability in risk factors, including SBP, modestly enhanced CVD risk discrimination.^13^ Given these conflicting findings, the present study aims to incorporate VVV BPV into a CVD risk prediction model and to evaluate its performance by comparing the discrimination and calibration of models using a single BP measurement versus those incorporating VVV BPV.

## Methods

### Study Design

This study was conducted in accordance with the principles outlined in the Transparent Reporting of a multivariable prediction model for Individual Prognosis or Diagnosis (TRIPOD)^19^ as presented in Supplementary Table 1. Data from the electronic practice-based research network (ePBRN) dataset, a resource compiled from a consortium of consenting GP clinics and secondary/tertiary hospitals in Southwestern Sydney, Australia, was utilised. Between 2006 and 2019, healthcare professionals, including GPs, hospital physicians, allied health professionals, and nurses, recorded patients’ information directly into the EHR system during routine clinical care. These records were later systematically extracted, de-identified, and linked using a third-party tool (GRHANITE™).

### Study Cohort

A landmark age of 55 years was used in this study; the study timeline is presented in Supplementary Figure 1. The landmark age serves as a reference point for risk prediction, based on VVV BP measurements collected up to that age. Landmark age models were constructed with a time origin at 55 years, incorporating prior BP records from eligible individuals. The inclusion criteria were as follows: 1) age 18-55 years with recorded BP measurements; 2) a minimum of five BP readings obtained across separate clinical encounters, with a minimum interval of two weeks between visits; and 3) no prior diagnosis of atherosclerotic CVD at the landmark age. Exclusion criteria included patients with missing gender or year of birth, or those without any recorded outpatient or hospital visits after their 55^th^ birthday.

The cohort development is depicted in Supplementary Figure 2. From an initial cohort of 20,757 patients with linked GP and hospital records, 6,397 were excluded because they had fewer than five BP records prior to age 55 years, 10,967 were excluded as they lacked outcome data, and 328 were excluded because outcome data were recorded before age 55 years. This resulted in a final analytic cohort of 3,065 patients.

### Power calculation

The power was estimated using standard formulas for Cox proportional hazards regression, assuming an expected event rate of 6.7% ^20^, and an alpha of 0.05. Power calculations were performed to detect the effect of each increase in each BPV metric. With an actual cohort size of 3,065, the study attained a power of 0.90.

### Outcome

The primary outcome was the first onset of CVD within the study cohort. CVD was delineated to encompass both fatal and non-fatal coronary heart disease (such as myocardial infarction and angina), cerebrovascular events (including stroke and transient ischemic attack), heart failure, and peripheral artery disease. The incidence of cases was determined by the first recorded CVD event documented in either the general practice or hospital datasets.

### Predictors

The following BP measurements were deemed biologically implausible and thus excluded: SBP <60 mmHg, SBP >250 mmHg, DBP <40 mmHg, and DBP > 140 mmHg. All BP values were measured during GP clinic visit. If there were multiple readings on the same day, the mean BP value was used in the analysis. In the basic model, the following variables were included: gender, smoking status, fasting blood glucose, body mass index (BMI), total cholesterol to high-density lipoprotein (TC/HDL) ratio, estimated glomerular filtration rate (eGFR), family history of CVD, presence of hypertension, diabetes mellitus, and a single SBP measurement obtained during one clinic visit. All predictor values were collected around the individual’s 55^th^ birthday. In the longitudinal model, the single BP measurement was replaced with a VVV BPV, assessed using SD and coefficient of variation (CV). SD values for systolic and diastolic BPV were categorised based on cut-offs from a previous study: 19 mmHg for systolic BPV and 11 mmHg for diastolic BPV. CV values were similarly categorised: 14% for systolic BPV and 12% for diastolic BPV.^21^

### Statistical analysis and model development

Multiple imputations using chained equations (MICE) were performed to address missing predictor data, generating 20 imputed datasets. Imputation and subsequent analyses were conducted in R version 4.4.0 using the MICE package.^22^ Cox proportional hazards model was fitted using the survival package version 3.7-0e.^23^

CVD predictive models were developed using Cox proportional hazards regression with 10-fold cross-validation on all imputed datasets. Model performance was evaluated for discrimination and calibration. Discrimination was assessed using Harrell’s C-index and the time-varying area under the curve (AUC) for annual CVD prediction up to five years. Calibration was evaluated using calibration slopes, and Brier scores. The Brier score was assessed annually over the five-year follow-up. Subgroup analysis was conducted to compare model performance between males and females using Harrell’s C-index, 5-year AUC, and calibration slope.

The time-to-event was defined as the interval between the participant’s 55th birthday and either the first diagnosis of CVD or the time of censoring. Participants without a CVD event were censored at their last recorded contact prior to 31 December 2019. Descriptive statistics are presented as means with standard deviations or as proportions, as appropriate.

## Results

### Characteristics of the study population

This study examined 3,065 patients, of whom 54.60% were men. Baseline characteristics are presented in Table 1. The average duration of BP monitoring was 6.44 ± 3.47 years. The mean SBP and DBP were 131.3 ± 21.5 mmHg and 80.4 ± 12.0 mmHg, respectively. Over a mean follow-up period of 4.26 ± 3.08 years, 231 patients (7.54%) developed CVD, while 2,836 patients (92.46%) either remained free of CVD or were censored. The prevalence of hypertension, diabetes mellitus, and dyslipidaemia was 31.40%, 6.88%, and 18.46%, respectively. The average number of BP measurements is 10.8 ± 8.6. The list of medications included in this study is presented in Supplementary Table 2.

**Table 1.** Baseline characteristics of the patients (before and after imputation of the dataset)

| Characteristics at Baseline | Data before imputation | Data after imputation |
| --- | --- | --- |
| Gender, n (%) |  | NA |
| Missing, n (%) | 0 (0) |  |
| Men | 1,673 (54.58) |  |
| Women | 1,392 (45.41) |  |
| Smoking status, n (%) |  |  |
| Missing, n (%) | 488 (15.92) | 0 (0) |
| Ex-smoker | 180 (5.87) | 224 (7.32) |
| Nonsmoker | 1,495 (48.78) | 1,781 (58.10) |
| Smoker | 902 (29.43) | 1,060 (34.60) |
| Body mass index, |  |  |
| Missing, n (%) | 1,819 (52.72) | 0 (0) |
| Mean ± SD | 29.54 ± 5.58 | 29.47 ± 11.88 |
| With diabetes mellitus, n (%) | 211 (6.88) | NA |
| With hypertension, n (%) | 963 (31.41) | NA |
| With dyslipidaemia, n (%) | 566 (18.46) | NA |
| Fasting blood glucose (mmol/L) |  |  |
| Missing, n (%) | 2,051 (66.91) | NA |
| Mean ± SD | 5.52 ± 1.61 | 5.43 ± 1.35 |
| Total cholesterol/HDL ratio |  |  |
| Missing, n (%) | 2,101 (68.54) | NA |
| Mean ± SD | 3.94 ± 1.13 | 4.00 ± 1.39 |
| eGFR (mL/min/1.73m<sup>2</sup>) |  |  |
| Missing, n (%) | 1,372 (44.76) | NA |
| Mean ± SD | 84.47 ± 9.25 | 83.84 ± 9.67 |
| Systolic blood pressure (mmHg),<br>mean ± SD | 131.34 ± 21.52 | NA |
| Diastolic blood pressure (mmHg),<br>mean ± SD | 80.37 ± 12.00 | NA |
| Anti-hypertensive use, n (%) | 874 (28.51) | NA |
| **Anti-diabetes medication use, n (%)** | 108 (3.52) | NA |
| Anti-dyslipidaemia use, n (%) | 516 (16.83) | NA |
| Anti-coagulant use, n (%) | 52 (1.70) | NA |

### Model performance comparison

The comparison of time-varying AUC is illustrated in Figure 1. Detailed values for each model are presented in Supplementary Table 1. For SBP, the time-varying AUCs of VVV BPV measured by SD and CV followed a similar pattern over the five-year period. A slight increase was observed between years two and three, with the highest five-year CVD prediction values reaching 0.852 (95% CI: 0.820 – 0.885) for SD and 0.856 (95% CI: 0.824 – 0.888) for CV. In contrast, the model based on a single SBP measurement exhibited markedly lower discriminative ability compared to the visit-to-visit BPV, with a five-year AUC of 0.757 (95% CI: 0.700–0.815). When single SBP and VVV BPV were combined in a model, the time-varying AUC did not differ from the model that utilised VVV BPV alone. For a five-year period, the time-varying AUC was 0.850 (95% CI: 0.816 – 0.884) for the combination of single SBP and SD and 0.853 (95% CI: 0.820 – 0.887) for the combination of single SBP and CV.

**Figure 1.**
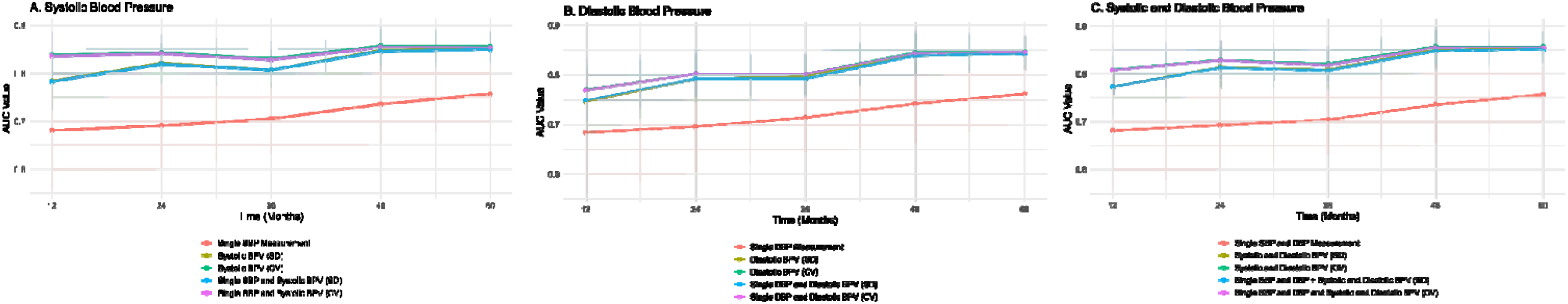
The comparison of time-varying area under the curve cardiovascular disease risk prediction models using single BP measurements, visit-to-visit BPV, assessed by SD and CV. BP, blood pressure; BPV, blood pressure variability; DBP, diastolic blood pressure; SBP, systolic blood pressure; SD, standard deviation; CV, coefficient of variation.

The analysis using DBP yielded slightly higher time-varying AUC results compared with SBP. Across the five-year follow-up, the time-varying AUCs for VVV BPV, evaluated using SD and CV, showed a consistent upward trend. These values are significantly higher compared to the model with single DBP only. When single DBP and VVV BPV were combined in a model, the time-varying AUC did not differ from that of the model utilising VVV BPV alone.

When both SBP and DBP were considered, model performance improved slightly compared to using DBP alone. The time-varying AUC for predicting CVD demonstrated a consistent increase over time. The overall five-year AUC values were comparable between the models using SD and CV. When both single measurements of SBP and DBP and their VVV BPV were combined in a model, the time-varying AUC did not differ from the model that used the combination of SBP or DBP VVV BPV alone.

The evaluation of overall model performance using Harrell’s C-index, Brier score, and calibration slope is summarised in Table 2. The longitudinal model incorporating VVV BPV, demonstrated superior discriminatory ability. Models including SD and CV consistently exhibited significantly higher Harrell’s C-index values compared with models using only a single BP measurement. For the SBP model, the C-index improved from 0.716 (95% CI: 0.658 – 0.775) with a single BP measurement to 0.833 (95% CI: 0.804 – 0.862) with SD and 0.837 (95% CI: 0.810 – 0.864) with CV. Similarly, for DBP, the C-index increased from 0.713 (95% CI: 0.655 – 0.772) to 0.820 (95% CI: 0.789 – 0.851) with SD and 0.825 (95% CI: 0.794 – 0.856) with CV. Importantly, the model combining both SBP and DBP single measurements yielded a similar C-index with SBP only, that is, 0.716 (95% CI: 0.657 – 0.774), while the inclusion of SD and CV improved it to 0.834 (95% CI: 0.804 – 0.863) and 0.839 (95% CI: 0.811 – 0.866), respectively. When both single measurements of SBP and DBP and their VVV BPV were combined in a model, Harrel’s C-Index did not differ from the model that used the combination of SBP and DBP VVV BPV alone; they are 0.833 (95% CI: 0.803 – 0.863) and 0.838 (95% CI: 0.810 – 0.866).

The average Brier scores were consistent across models that used VVV BPV and those that used only a single BP measurement, indicating no enhancement in the Brier score when replacing single SBP and/or DBP measurements with VVV BPV or when combining VVV BPV with either single SBP or DBP. The details of the time-varying Brier score are depicted in Supplementary Table 3.

**Table 2.**
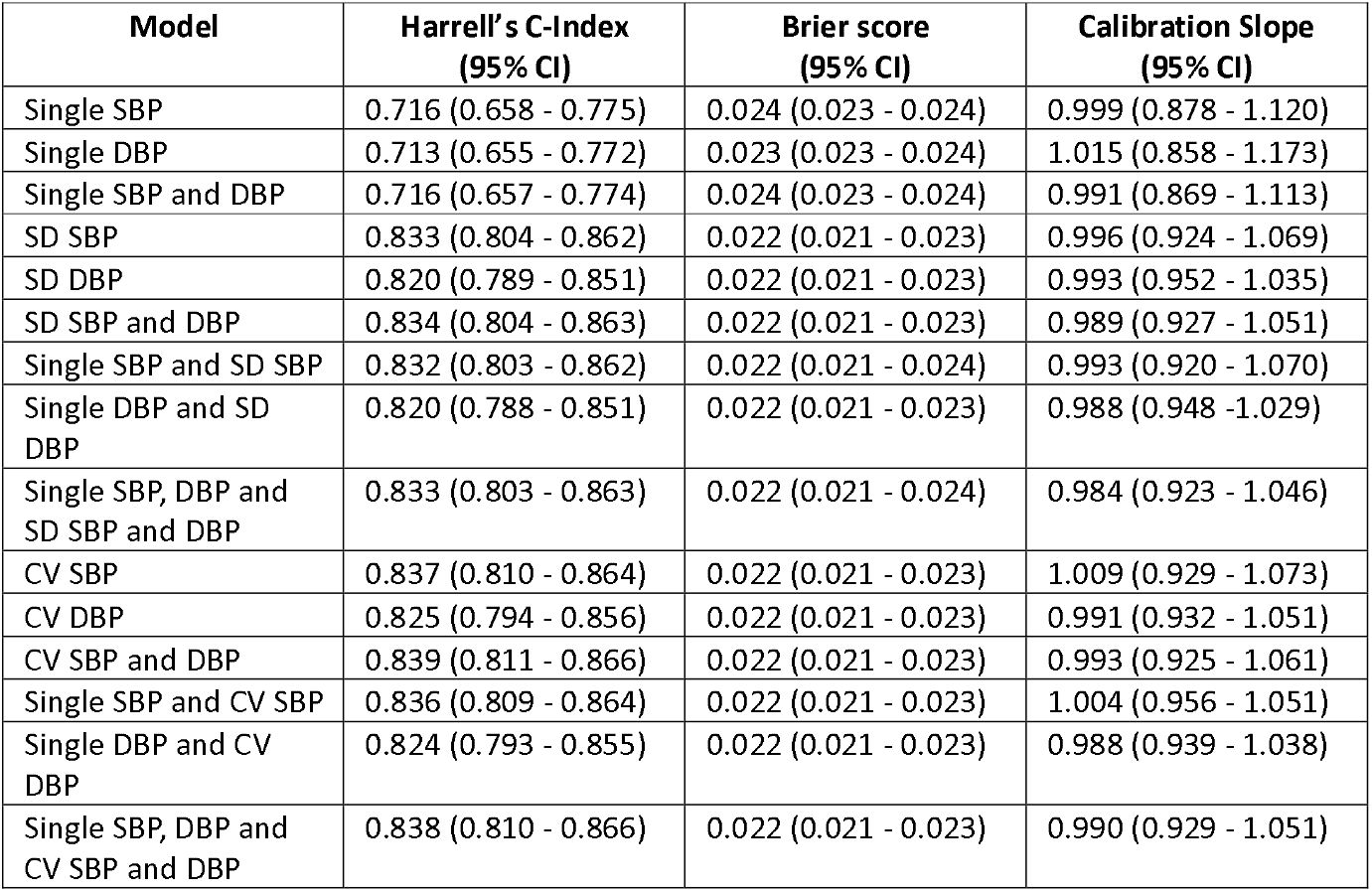
Harrell C-index, Brier score, and calibration slope for cardiovascular disease risk prediction.

**Table 3.** Net Reclassification Index for cardiovascular disease risk prediction.

| Model | Net Reclassification Index (NRI) |  |  |
| --- | --- | --- | --- |
|  | NRI + (95% CI) | NRI - (95% CI) | Overall |
| **SBP** |  |  |  |
| Single SBP | Reference | Reference | Reference |
| SD SBP | 0.167 (0.063 – 0.272) | 0.273 (0.219 – 0.327) | 0.440 (0.319 – 0.562) |
| Single SBP and SD SBP | 0.169 (0.064 – 0.273) | 0.273 (0.219 – 0.326) | 0.441 (0.318 – 0.566) |
| CV SBP | 0.159 (0.053 – 0.265) | 0.271 (0.217 – 0.326) | 0.431 (0.308 – 0.554) |
| Single SBP and CV SBP | 0.163 (0.0581 – 0.266) | 0.271 (0.217 – 0.325) | 0.433 (0.311 – 0.557) |
| **DBP** |  |  |  |
| Single DBP | Reference | Reference | Reference |
| SD DBP | 0.135 (0.041 – 0.230) | 0.268 (0.207 – 0.330) | 0.405 (0.281 – 0.528) |
| Single DBP and SD DBP | 0.134 (0.038 – 0.231) | 0.270 (0.208 – 0.330) | 0.404 (0.279 – 0.529) |
| CV DBP | 0.183 (0.084 – 0.281) | 0.223 (0.165 – 0.280) | 0.406 (0.286 – 0.526) |
| Single DBP and CV SBP | 0.183 (0.083 – 0.281) | 0.222 (0.165 – 0.281) | 0.405 (0.285 – 0.525) |
| **SBP and DBP** |  |  |  |
| Single SBP and DBP | Reference | Reference | Reference |
| SD SBP and DBP | 0.157 (0.048 – 0.266) | 0.277 (0.223 – 0.331) | 0.434 (0.304 – 0.564) |
| Single SBP, DBP and SD SBP and DBP | 0.156 (0.045 – 0.268) | 0.277 (0.224 – 0.330) | 0.434 (0.301 – 0.567) |
| CV SBP and DBP | 0.182 (0.066 – 0.297) | 0.254 (0.201 – 0.308) | 0.436 (0.302 – 0.571) |
| Single SBP, DBP and CV SBP and DBP | 0.186 (0.070 – 0.304) | 0.255 (0.202 – 0.309) | 0.442 (0.302 – 0.582) |

Calibration slopes also remained consistent across models that included VVV BPV and those that used only a single BP measurement. The values were closer to the ideal value of 1, with confidence intervals encompassing one, suggesting that these models neither over- nor under-predicted events. Subgroup analysis comparing model performance based on Harrell’s C-index, 5-year AUC, and calibration slope between males and females is shown in Supplementary Table 4. The model performance in both males and females showed similar Harrell’s C index, 5-year AUC, and calibration slope when using VVV BPV. However, the calibration slope for the male subgroup, based on a single SBP/DBP measurement, suggests the model underpredicts risk in males.

## Discussion

This study demonstrated that substituting a single BP measurement with VVV BPV, measured using SD and CV, significantly enhanced the predictive performance of CVD risk models. The incorporation of VVV BPV notably improved model calibration, accuracy, and discrimination. Interestingly, the combination of a single BP measurement with VVV BPV did not further enhance model performance. The parameters of model performance remained consistent with those of the model replacing a single BP measurement with VVV BPV. The consistent improvement observed across models utilising VVV BPV of SBP and/or DBP indicates that BPV serves as a robust and reliable predictor of CVD risk.

The comparable reliability of SD and CV across models underscores the clinical significance of BPV as a dynamic biomarker of CVD risk. In this study, the cut-off values for SD and CV were derived from our previous study ^21^, with thresholds of SD for SBP >19 mmHg, SD for DBP >11 mmHg, CV for SBP >14%, and CV for DBP >11%. Although SD and CV produced comparable results, models using CV, particularly in DBP, demonstrated slightly higher Harrell’s C-Index and time-varying AUC. CV, being a dispersion measure normalised to the mean, provides advantages in populations with heterogeneous baseline BP levels. This observation aligns with previous studies showing that CV outperformed both SD and average real variability (ARV) ^16^.

Models incorporating diastolic BPV showed slightly better predictive performance than those using systolic BPV only. This finding is consistent with a previous systematic review and meta-analysis, which showed that diastolic BPV was associated with a higher pooled hazard ratio for CVD events ^9^. The superior performance of DBP variability may reflect its sensitivity to arterial stiffness, disrupted coronary perfusion, and impaired endothelial function ^24^. Furthermore, combining both SBP and DBP variability yielded the highest predictive metrics, suggesting that the combined assessment offers a more comprehensive representation of vascular and hemodynamic alterations underlying CVD risk.

### Clinical, health policy, and future research implications

Integrating dynamic VVV BPV calculation into the EHR, along with an integrated dynamic CVD risk score, would enhance CVD risk assessment within GP clinics and support primary prevention efforts for CVD. The utilisation of VVV BPV may serve as an important independent predictor of CVD risk in this group ^25^. Unlike a single static BP measurement, BPV likely captures the early signs of vascular aging and alterations in the cardiac structure that occur before the onset of CVD. Prior studies have shown that VVV BPV beginning in childhood or early adulthood is associated with vascular aging, increased carotid intima-media thickness, and cardiac remodelling in midlife ^26–28^.

Enhancing awareness among the public and clinicians about the importance of longitudinal BP measurements could improve the management of hypertension in the population. Previous research shows that young to middle–aged adults generally have lower awareness of hypertension and CVD events compared to older populations.^29,30^ Our dataset also indicates that the younger cohort tends to have fewer BP records in GP clinics. BP measurement should be included in the assessment for any visit, regardless of whether it’s related to cardiovascular symptoms or the age of the patients. This approach would increase the number of BP recordings in GP clinics, enabling direct integration with the dynamic CVD risk score embedded in the EHR. Incorporating VVV BPV into CVD risk scores could support a more personalised and dynamic approach to CVD prevention, especially for adults under 55 years old.

Our earlier study found that, after adjusting for medication use, the hazard ratio linking VVV BPV to CVD remained largely unchanged.^21^ This suggests that VVV BPV is an independent risk factor for CVD. Moreover, previous studies indicate that calcium channel blockers are more effective in reducing VVV BPV than other antihypertensive classes, such as beta blockers, angiotensin receptor blockers, and diuretics.^31^ Additionally, research has shown that VVV BPV is associated with CVD outcomes in patients with optimal SBP level (<140 mmHg), encompassing patients with and without anti-hypertensive therapy.^32^ Thus, VVV BPV can serve as a predictor for estimating residual CVD risk after antihypertensive treatment and achieving optimal BP targets. Furthermore, higher systolic VVV BPV has been linked to poor adherence to antihypertensive medication.^33^ Since poor medication adherence is a challenging risk factor to measure, incorporating VVV BPV into CVD risk scores could also help account for hypertension management and medication adherence.

This study exclusively examines individuals younger than 55 years, with BP records and forecasts their risk of CVD after turning 55. Future research using different landmark ages, especially those at younger ages, is needed to enhance the model’s robustness across the broader population. Additionally, assessing the net reclassification index between the Australian CVD risk score and the CVD risk score incorporating VVV BPV would further enhance the model’s utility.

### Strengths and Limitations

This study utilised longitudinal blood pressure data derived from EHRs in general practice, linked with hospital data, thereby offering valuable insights into real-world clinical practice. However, as the data were obtained from EHRs, certain variables, including risk factors, laboratory results and body mass index, had missing values that necessitated multiple imputation to include those variables in the model for predicting CVD.

We assessed VVV BPV for each patient prior to their reaching 55 years of age to establish a clear timeline between exposure and outcome, ensuring that the BPV evaluation occurred before the onset of CVD. However, this method may have excluded those who develop early-onset CVD, which could lead to an under-appreciation of CVD risk among younger individuals at high risk. Additionally, this study only calculated the risk of CVD for individuals aged 55 years and older, as it had a limited number of samples with consistent collection of BP records in primary care prior to the age of 55 years. As a result, the proportion of smokers within this study is also elevated, given that having at least five blood pressure measurements in this dataset indicates that these patients typically have more chronic health issues. This reality indicates that the findings of this study may not be applicable to the general population. Further research is necessary to validate these results externally.

## Conclusions

This study developed a model for CVD risk estimation using VVV BPV instead of a single blood pressure measurement. Replacing a single BP measure with VVV BPV significantly enhanced the model’s predictive accuracy. These results highlight the importance of regular BP monitoring and underscore the need to incorporate VVV BPV-based risk scores into GP EHR systems to enable early CVD risk identification and prevention.

## Competing Interests

JR has received honorarium from Pfizer and Merck Sharpe & Dohme for providing advice, chairing and speaking at clinician educational events.

## Data availability statement

The corresponding author will make the code and data available with valid reasons and approval when required.

## Funding

This study was funded by the Australian National Health and Medical Research Council (Grant Number: GNT1192469). JJ also acknowledges the funding support received through the Research Technology Services at UNSW Sydney, Google Cloud Research (Award Number: GCP19980904), and the NVIDIA Academic Hardware grant programs.

## Role of the Funder/Sponsor

The sponsors had no role in the design and conduct of the study; collection, management, analysis, and interpretation of the data; preparation, review, or approval of the manuscript; and decision to submit the manuscript for publication.

## Acknowledgements

We want to thank the ePBRN Primary Care Health Informatics Working Group of the Secure Research Environment for Digital Health (SREDH) Consortium (www.sredhconsortium.org, accessed on 7 October 2024) for their assistance with access to the ePBRN dataset to investigate the findings from this review.

## Authors’ contributions (CReDIT statement)

Conceptualization: BJ, JJ, JR, ML, NN, STL; Formal Analysis: ML; Investigation: JJ, ML; Methodology: BJ, JJ, JR, ML, NN, STL; Supervision: BJ, JJ, JR, NN, STL; Writing – original draft: ML; Writing – review & editing: BJ, JJ, JR, ML, STL, NN; All authors read and approved the final manuscript.

## Patient consent for publication

Not applicable.

## Ethics approval

The ethics of this study has been approved by Human Research Ethics Committee of the University of New South Wales, Sydney, Australia (approval number: HC230072).

## Patient and public involvement

This study relied on secondary data from primary care and hospitals; patients were not involved in the development of the model.

## References

1. Brown S, Banks E, Woodward M, Raffoul N, Jennings G, Paige E. Evidence supporting the choice of a new cardiovascular risk equation for Australia. Medical Journal of Australia. 2023;219(4):173–186. doi:10.5694/mja2.52052

2. Gyldenkerne C, Mortensen MB, Kahlert J, et al. 10-Year Cardiovascular Risk in Patients With Newly Diagnosed Type 2 Diabetes Mellitus. Journal of the American College of Cardiology. 2023;82(16):1583–1594. doi:10.1016/j.jacc.2023.08.015

3. Paige E, AM EB, Zhang Y, et al. Development and calibration of the 2023 Australian cardiovascular disease risk prediction equations: a model updating study. Med J Aust. 2025;223(4). Accessed October 27, 2025. https://www.mja.com.au/journal/2025/223/4/development-and-calibration-2023-australian-cardiovascular-disease-risk

4. Pylypchuk R, Wells S, Kerr A, et al. Cardiovascular disease risk prediction equations in 400 000 primary care patients in New Zealand: a derivation and validation study. The Lancet. 2018;391(10133):1897–1907. doi:10.1016/S0140-6736(18)30664-0

5. SCORE2 working group and ESC Cardiovascular risk collaboration. SCORE2 risk prediction algorithms: new models to estimate 10-year risk of cardiovascular disease in Europe. European Heart Journal. 2021;42(25):2439–2454. doi:10.1093/eurheartj/ehab309

6. Zomer E, Owen A, Magliano DJ, Liew D, Reid C. Validation of two Framingham cardiovascular risk prediction algorithms in an Australian population: the ‘old’ versus the ‘new’ Framingham equation. Eur J Cardiovasc Prev Rehabil. 2011;18(1):115–120. doi:10.1097/HJR.0b013e32833ace24

7. Wang Z, Hoy WE. Is the Framingham coronary heart disease absolute risk function applicable to Aboriginal people? Med J Aust. 2005;182(2):66–69.

8. Diaz KM, Tanner RM, Falzon L, et al. Visit-to-Visit Variability of Blood Pressure and Cardiovascular Disease and All-Cause Mortality. Hypertension. 2014;64(5):965–982. doi:10.1161/HYPERTENSIONAHA.114.03903

9. Lukitasari M, Jonnagaddala J, Liaw ST, Jalaludin B. Visit-to-visit blood pressure variability and cardiovascular outcomes: a systematic review and dose-response meta-analysis. Eur J Prev Cardiol. Published online August 28, 2025. doi:10.1093/eurjpc/zwaf546

10. Saputra PBT, Lamara AD, Saputra ME, et al. Long-term systolic blood pressure variability independent of mean blood pressure is associated with mortality and cardiovascular events: A systematic review and meta-analysis. Current Problems in Cardiology. 2024;49(2):102343. doi:10.1016/j.cpcardiol.2023.102343

11. Wang J, Shi X, Ma C, et al. Visit-to-visit blood pressure variability is a risk factor for all-cause mortality and cardiovascular disease: a systematic review and meta-analysis. Journal of Hypertension. 2017;35(1):10. doi:10.1097/HJH.0000000000001159

12. McAlister FA, Lethebe BC, Leung AA, Padwal RS, Williamson T. Visit-to-visit blood pressure variability is common in primary care patients: Retrospective cohort study of 221,803 adults. PLOS ONE. 2021;16(4):e0248362. doi:10.1371/journal.pone.0248362

13. Xu Z, Arnold M, Sun L, et al. Incremental value of risk factor variability for cardiovascular risk prediction in individuals with type 2 diabetes: results from UK primary care electronic health records. International Journal of Epidemiology. 2022;51(6):1813–1823. doi:10.1093/ije/dyac140

14. Ribero VA. Development and validation of the CARE-DM model to predict the cardiovascular risk in older persons with type 2 diabetes.

15. Paige E, Barrett J, Stevens D, et al. Landmark Models for Optimizing the Use of Repeated Measurements of Risk Factors in Electronic Health Records to Predict Future Disease Risk. Am J Epidemiol. 2018;187(7):1530–1538. doi:10.1093/aje/kwy018

16. Li J, Qu T, Li Y, et al. Long-term blood pressure variability and risk of cardiovascular diseases in populations with different blood pressure status: an ambispective cohort study. Blood Pressure Monitoring. 2024;29(5):249. doi:10.1097/MBP.0000000000000712

17. Paige E, Barrett J, Pennells L, et al. Use of Repeated Blood Pressure and Cholesterol Measurements to Improve Cardiovascular Disease Risk Prediction: An Individual-Participant-Data Meta-Analysis. Am J Epidemiol. 2017;186(8):899–907. doi:10.1093/aje/kwx149

18. Hippisley-Cox J, Coupland C, Brindle P. Development and validation of QRISK3 risk prediction algorithms to estimate future risk of cardiovascular disease: prospective cohort study. BMJ. 2017;357:j2099. doi:10.1136/bmj.j2099

19. Collins GS, Moons KGM, Dhiman P, et al. TRIPOD+AI statement: updated guidance for reporting clinical prediction models that use regression or machine learning methods. Published online April 16, 2024. doi:10.1136/bmj-2023-078378

20. Australian Bureau of Statistics. Causes of Death, Australia.; 2023. https://www.abs.gov.au/statistics/health/causes-death/causes-death-australia/latest-release

21. Lukitasari M, Argha R, Siaw-Teng L, Jalaludin B, Rhee J, Jonnagaddala J. Defining cut-offs for visit-to-visit blood pressure variability to predict cardiovascular disease in primary care patients. Manuscript submitted for publication. Published online 2025.

22. Buuren SV, Groothuis-Oudshoorn K. micel1: Multivariate Imputation by Chained Equations in R. J Stat Soft. 2011;45(3). doi:10.18637/jss.v045.i03

23. Therneau T. A package for survival analysis in R. Published online 2024. https://cran.r-project.org/web/packages/survival/index.html

24. Yu J, Song Q, Bai J, et al. Visit-to-Visit Blood Pressure Variability and Cardiovascular Outcomes in Patients Receiving Intensive Versus Standard Blood Pressure Control: Insights From the STEP Trial. Hypertension. 2023;80(7):1507–1516. doi:10.1161/HYPERTENSIONAHA.122.20376

25. Yano Y, Reis JP, Lewis CE, et al. Association of Blood Pressure Patterns in Young Adulthood With Cardiovascular Disease and Mortality in Middle Age. JAMA Cardiol. 2020;5(4):382–389. doi:10.1001/jamacardio.2019.5682

26. Nwabuo CC, Yano Y, Moreira HT, et al. Long-Term Blood Pressure Variability in Young Adulthood and Coronary Artery Calcium and Carotid Intima-Media Thickness in Midlife: The CARDIA Study. Hypertension. 2020;76(2):404–409. doi:10.1161/HYPERTENSIONAHA.120.15394

27. Nwabuo CC, Yano Y, Moreira HT, et al. Association Between Visit-to-Visit Blood Pressure Variability in Early Adulthood and Myocardial Structure and Function in Later Life. JAMA Cardiol. 2020;5(7):795–801. doi:10.1001/jamacardio.2020.0799

28. Wu GJ, Si AM, Wang Y, et al. Associations of ultra long-term visit-to-visit blood pressure variability, since childhood with vascular aging in midlife: a 30-year prospective cohort study. Journal of Hypertension. 2024;42(11):1948–1957. doi:10.1097/HJH.0000000000003819

29. Allen N, Wilkins JT. The Urgent Need to Refocus Cardiovascular Disease Prevention Efforts on Young Adults. JAMA. 2023;329(11):886–887. doi:10.1001/jama.2023.2308

30. Steckelmacher J, Faconti L, Gupta A. Beyond the Tip of the Iceberg: Rethinking Hypertension Screening in Young Adults. Journal of the American Heart Association. 2025;14(14):e042877. doi:10.1161/JAHA.125.042877

31. Nardin C, Rattazzi M, Pauletto P. Blood Pressure Variability and Therapeutic Implications in Hypertension and Cardiovascular Diseases. High Blood Press Cardiovasc Prev. 2019;26(5):353–359. doi:10.1007/s40292-019-00339-z

32. Liu M, Chen X, Zhang S, et al. Assessment of Visit-to-Visit Blood Pressure Variability in Adults With Optimal Blood Pressure: A New Player in the Evaluation of Residual Cardiovascular Risk? J Am Heart Assoc. 2022;11(9):e022716. doi:10.1161/JAHA.121.022716

33. Muntner P, Levitan EB, Joyce C, et al. Association Between Antihypertensive Medication Adherence and Visit-to-Visit Variability of Blood Pressure. The Journal of Clinical Hypertension. 2013;15(2):112–117. doi:10.1111/jch.12037

